# Multi-task artificial intelligence annotation of echocardiographic images: a retrospective multi-cohort study

**DOI:** 10.64898/2026.06.23.26356383

**Authors:** Yuki Sahashi, David Choi, Hirotaka Ieki, Milos Vukadinovic, Meenal Rawlani, Bryan He, Alan C. Kwan, Susan Cheng, David Ouyang

## Abstract

**Background:** A comprehensive transthoracic echocardiogram involves the assessment of over 70 parameters, placing a substantial burden on sonographers and physicians for manual annotation with considerable inter-observer variability. Prior open-source segmentation models have largely addressed 2D B-mode ventricular function, leaving a gap in the spectral Doppler and atrial measurements required for valvular and diastolic assessment such as velocity-time integral (VTI) and atrial chamber size.

**Methods:** In this retrospective multi-cohort study, we developed EchoNet-Segmentation, comprehensive task-specific deep learning segmentation models for left and right atrial area and VTI Doppler measurements. Training used 186,712 sonographer-annotated images from 93,978 studies (56,855 patients) at Cedars-Sinai Medical Center (CSMC). Performance was evaluated on a held-out CSMC test set, a CSMC temporal split, an external Kaiser Permanente Northern California cohort, and the public MIMIC-Echo dataset.

**Findings:** On the CSMC held-out test set, our AI models showed strong agreement with sonographer measurements, with R² of 0.817–0.882 and mean absolute error (MAE) of 1.13–3.80 cm for automated VTI measurements, and R² of 0.675–0.747 and MAE of 2.48–2.52 cm² for left and right atrial area segmentation. Performance was consistently confirmed on the CSMC temporal split (VTI: R² 0.606–0.866, atrial area: R² 0.694–0.705) and on the KPNC external cohort (VTI: R² 0.575–0.859, atrial area: R² 0.803–0.876), on the MIMIC-Echo dataset. Robustness was demonstrated on a different vendor’s machines and across subgroups. EchoNet-Segmentation outperformed an open-source medical image foundation model with bounding-box, point prompt configurations on R², MAE, and Dice score on both held-out test dataset and MIMIC apical four-chamber data.

**Interpretation:** EchoNet-Segmentation is the first open-source framework that delivers accurate, generalizable automated measurement across several key routine echocardiographic parameters, supporting end-to-end automation of clinically important echocardiographic assessments. Public release of model weights, code, and demonstration tools can facilitate reproducibility, research use and clinical deployment.

**Funding:** Funding Statement: This work was supported by NIH NHLBI grants R00HL157421, R01HL173526, and R01HL173487 to D.O.

**Research in context:** *Evidence before this study:* We searched PubMed and arXiv from database on April 1, 2026, for studies of deep learning-based segmentation of echocardiographic images, using the terms (“echocardiography” OR “echocardiogram”) AND (“deep learning” OR “artificial intelligence”) AND (“segmentation” OR “measurement”). Prior work has demonstrated automated segmentation of cardiac chambers and left ventricular ejection fraction estimation, and a small number of studies have reported deep learning models for velocity-time integral (VTI) or atrial size measurement. However, openly available models and code remain largely restricted to left ventricular structures, ejection fraction, and wall thickness, and commercial tools remain proprietary. To our knowledge, no open-source framework has comprehensively addressed VTI measurements across multiple Doppler views together with atrial chamber size in a single, reproducible toolkit, and existing models have not been systematically benchmarked against general-purpose medical-image foundation models on echocardiographic tasks.

*Added value of this study:* We developed and validated EchoNet-Segmentation, a suite of task-specific deep learning models for several clinically important echocardiographic parameters: left and right atrial area and five VTI measurements (aortic valve, mitral valve, left ventricular outflow tract, right ventricular outflow tract, and pulmonary valve). The models were trained on the largest real-world collection of sonographer-annotated echocardiograms reported to date (186,712 images from 56,855 patients) in an academic center in the United States and showed strong agreement with sonographer measurements on a held-out internal test set, a temporal split cohort, an external cohort from a different health system, and a publicly available cohort recorded on a different vendor’s ultrasound machines. EchoNet-Segmentation outperformed the publicly released medical-image foundation model (MedSAM2) on cardiac chamber segmentation across both internal and public dataset benchmarks. All model weights, training and inference code, demonstration tools, and the manual segmentation masks used for the public benchmark are openly released.

*Implications of all the available evidence:* EchoNet-Segmentation enables end-to-end automation of routine transthoracic echocardiographic measurements with previously released open-source models. By openly releasing model weights, training code, and benchmark data, this work provides a reproducible foundation that the broader research and clinical community can build on, fine-tune for specific populations or imaging protocols, and integrate into clinical workflows. Prospective validation and randomized studies will be needed to define the impact of automated measurement on diagnostic accuracy, workflow efficiency, and clinical outcomes.

## Introduction

Echocardiography is the most widely used cardiac imaging modality, playing a central role in the screening, surveillance, severity assessment, and preoperative evaluation of cardiovascular disease^1,2^. The volume of echocardiographic examinations continues to grow substantially^3^, and accurate measurement is essential for appropriate clinical decision-making. A comprehensive echocardiographic examination involves the assessment of over 70 distinct parameters, each carrying clinical significance^2,4^. Given the breadth of these measurements, manual segmentation places a significant burden on clinical practice and is subject to considerable inter-observer variability^5^.

The advent of deep learning in computer vision has enabled automated image analysis across a broad spectrum of medical imaging applications^6–11^. In echocardiography, task-specific models have been proposed for wall thickness measurement^12–14^, ejection fraction estimation^15,16^, and selected Doppler measurements^12,17^. More recently, vision-language models have further advanced the field by enabling automated report generation^8,18^. Despite these advances, comprehensive segmentation-based measurement covering the full breadth of clinically relevant parameters has not yet been achieved. Commercially available tools remain proprietary, and existing open-source models are limited in scope^19,20^. Prior open-source efforts have largely addressed 2D B-mode ventricular function, leaving a gap such as the spectral Doppler and atrial structures for valvular, shunt and diastolic assessment. Accurate VTI measurement is essential for aortic valve area calculation by the continuity equation, the standard method for grading aortic stenosis severity^21^, as well as for valvular disease management, diastolic function assessment^22^, and shunt volume quantification. Similarly, atrial volume is critical for the diagnosis of heart failure with preserved ejection fraction and estimation of left atrial pressure^22,23^. These parameters are also among those with the highest inter-observer variability, making them particularly well-suited targets for automation^5^.

We previously developed the first open-source deep learning framework for automated measurement of 18 echocardiographic parameters^12^. To advance precision medicine in cardiovascular imaging, we now extend this work by developing and validating open-source area segmentation models for additional clinically important parameters, including VTI measurements, left atrial volume, and right atrial volume. To support the broader research community, we publicly release model weights, tutorials, and code for fine-tuning to facilitate further development and clinical deployment.

## Methods

### Data collection and procedures

For the derivation cohort, we identified all resting transthoracic echocardiography studies performed at Cedars-Sinai Medical Center (CSMC) between November 2009 and June 2022, along with the corresponding sonographer and cardiologist annotations. A total of 93,978 studies from 56,855 patients who received a clinical transthoracic echocardiogram were included (**Figure 1**). No exclusion criteria were applied based on patient characteristics or machine vendors. Imaging data were stored in Digital Imaging and Communications in Medicine (DICOM) format and annotated by expert sonographers and cardiologists as part of routine clinical practice.

**Figure 1:**
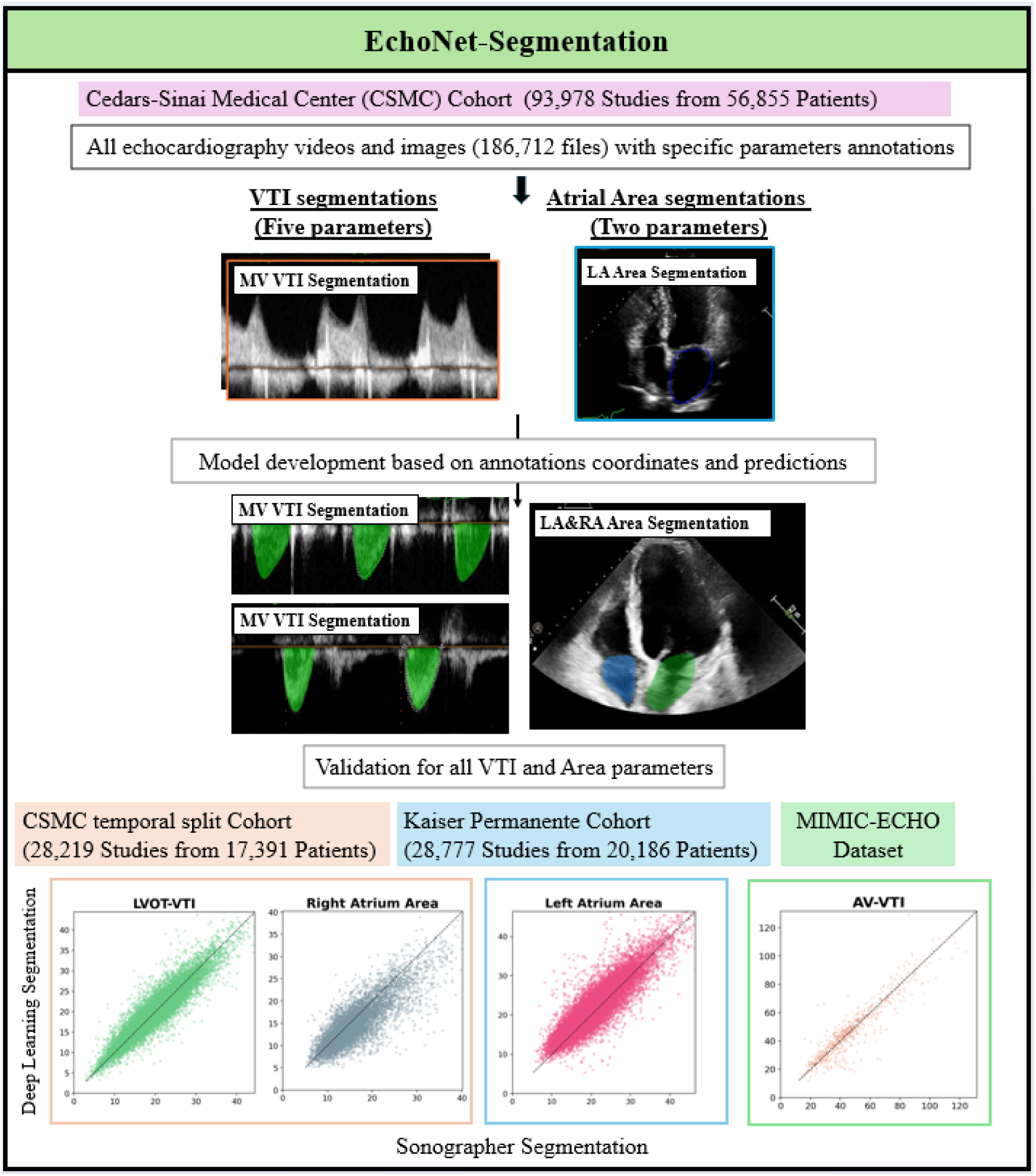
Overview of the study pipeline.

The target measurements in this study were grouped into two categories: velocity-time integral (VTI) measurements and atrial area measurements. The VTI group included aortic valve VTI (AV VTI), mitral valve VTI (MV VTI), left ventricular outflow tract VTI (LVOT VTI), right ventricular outflow tract VTI (RVOT VTI), and pulmonary valve VTI (PV VTI). The atrial area group included left atrial area (LA area) and right atrial area (RA area). For the held-out test dataset, image-level analysis was performed to directly compare the AI model output with the sonographer measurement in the same image.

We evaluated the model performance on 28,777 echocardiography studies at Kaiser Permanente Northern California (KPNC) between January 2022 and April 2022 as external validation, and on 28,219 studies between June 2022 and November 2024 at CSMC as a temporal split validation dataset. In the end-to-end approach, automated parameter measurements corresponding to each view were performed following the view classification. To assess model robustness across different vendors and data formats, we additionally performed inference on a subsample of the publicly available MIMIC-Echo dataset^24^. As the MIMIC-Echo dataset contained measurement reports only for AV VTI and LVOT VTI, we limited the VTI evaluation to these two VTI measurements. For atrial measurements, we derived the reference values from the left atrial volume and right atrium length instead of both atrial areas due to the lack of those ground-truth value. Left atrium volume were calculated using the area-length method at end-systole per the guideline^2^. Echocardiography studies in the CSMC cohort and the KPNC external validation cohort were performed using Philips ultrasound machines (Philips EPIQ 5C, 7C, iE 33 or EPIQ CVx), and those in the MIMIC-Echo dataset were conducted using GE Healthcare ultrasound systems (Vivid E90, Vivid E95, and Vivid S7).Approval for this study was obtained from the Cedars-Sinai Medical Center and Kaiser Permanente Northern California Institutional Review Boards, and the requirement for informed consent was waived for retrospective data analysis.

### Model Development

For the atrial area segmentation models, we used images with corresponding sonographer-annotated coordinates for area measurements to generate corresponding segmentation masks. Input images were resized to 480 (height) by 640 (width) prior to training. Separate task-specific models were developed for LA area and RA area segmentation. For VTI measurements, we developed two separate models: a YOLO-based detection model and a segmentation model. Since each VTI image typically contains several Doppler wave segments but sonographer annotations are available for only one segment per image, we used only the annotated segment for training both models. The detection model (YOLOv11n^25^) was trained to localize each individual segment, with detection performance evaluated using mAP@50 and mAP@50-95 (training details provided in **Supplemental Methods**). Using the cropped VTI envelopes by the detection model, the segmentation model was trained to predict the area under the Doppler wave segments for VTI calculation (**Figure 2**).

**Figure 2:**
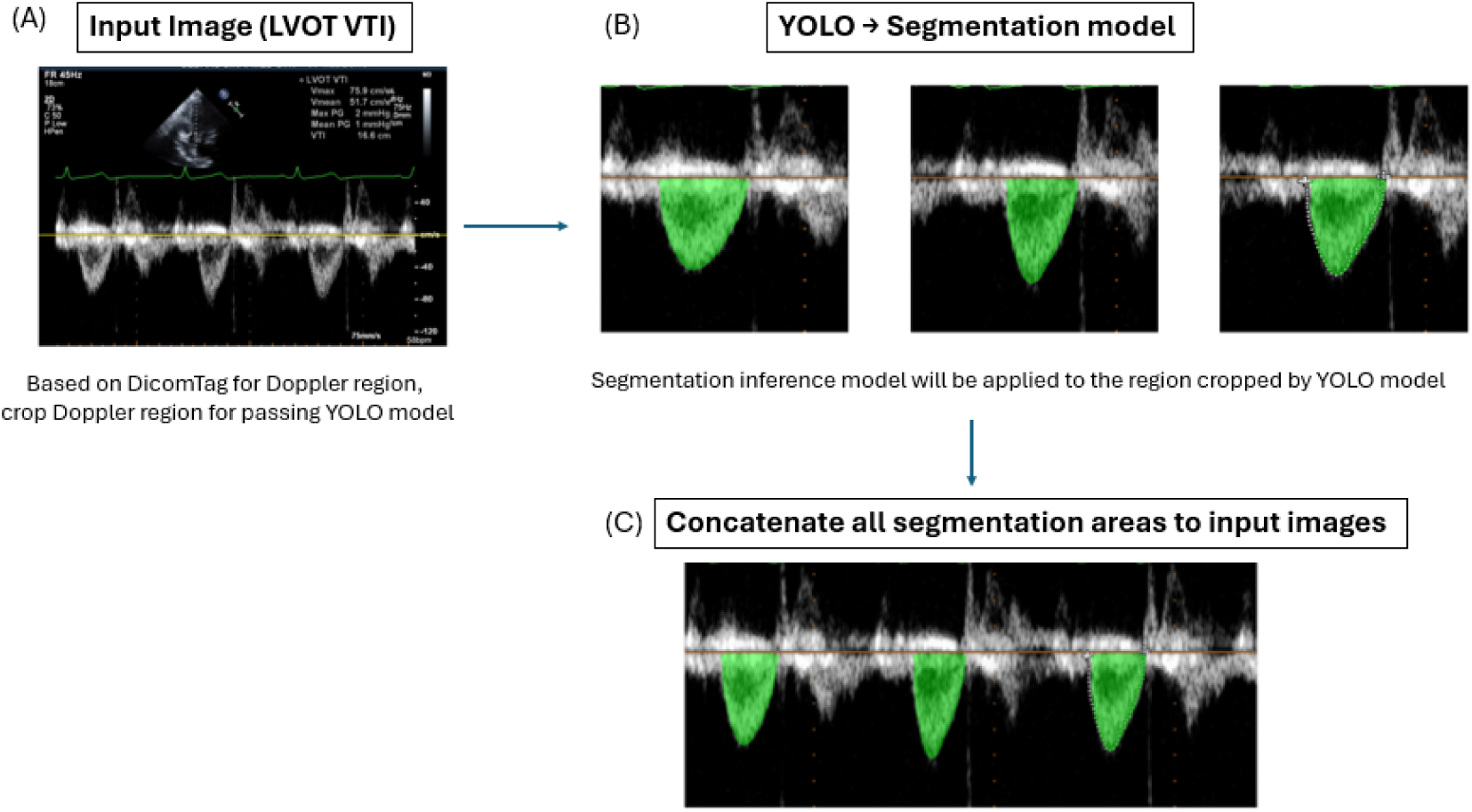
End-to-end pipeline for automated VTI measurement. Schematic of the two-stage deep learning pipeline for automated velocity-time integral (VTI) measurement. In the first stage, a YOLOv11n object-detection model identifies and localizes individual Doppler envelope segments (A) within each spectral Doppler image, generating bounding boxes around each candidate VTI waveform (B). In the second stage, each detected segment is cropped and passed to a DeepLabv3 segmentation model, which predicts the pixel-level area under the Doppler curve corresponding to a single cardiac cycle (C). The segmented area is then converted to a clinical VTI value (cm) using the calibration metadata embedded in the DICOM image. AI-derived segmentations are shown in green.

For both the atrial area and VTI segmentation models, we used a DeepLabv3 architecture^26^ trained with a binary cross-entropy with logits loss. The CSMC dataset was split by patients at a ratio of 80:10:10 for training, validation, and held-out testing. Models were trained using the Adam optimizer with a learning rate of 0.001, a batch size of 48, and up to 150 epochs with early stopping after 10 epochs of no improvement in validation loss. The weights from the epoch with the lowest validation loss were used for evaluation on the held-out test and external validation datasets. Dice score was used as the training-time evaluation metric for the segmentation model. The atrial area (cm^2^) for the area models and the calculated VTI (cm) for the VTI models were used as the primary clinical evaluation metrics. For the development cohort, all performance metrics were calculated using held-out test data that was not involved in model training.

### Comparison with publicly available segmentation model

We compared EchoNet-Segmentation with MedSAM2, a publicly released promptable foundation model built on the SAM2 backbone^27^. The released echocardiography checkpoint (“*MedSAM2_US_Heart.pt*”) was developed to segment four chambers and myocardium in apical four chambers using the RVENet dataset^28^ (3,583 apical echocardiographic videos from 831 patients) and human-in-the-loop annotation, enabling chamber and myocardial segmentation when supplied with either a box prompt or a point prompt at inference time. We first applied MedSAM2 with box and point prompts to the CSMC held-out test set (LA area and RA area subset). Inference with MedSAM2 weights without any prompts was also compared as zero-shot prediction. Then, we applied MedSAM2 with the echocardiography weight to randomly sampled MIMIC-ECHO apical four chamber dataset (n=300). For this experiment, ground-truth segmentations and bounding-box prompts were generated manually for each image by independent cardiologists who were not involved in model development, and the point prompt was defined as the centroid of the bounding box. These MedSAM2 predictions for LA area and RA area were compared against EchoNet-Segmentation predictions. Since the MedSAM2 checkpoint for echocardiography is trained only on the apical view for cardiac chambers, we did not perform a comparison with EchoNet-Segmentation for the VTI segmentation.

### Statistical analysis

Mean absolute error (MAE) and coefficient of determination (R²) between clinically measured parameters and parameters predicted by the deep-learning model were calculated. Bland-Altman plots, where the average of the two measurements was plotted against the difference, were used to assess the agreement between the actual and predicted measurements. Stratified analyses were performed based on the following patient characteristics: age (<65 or ≥65 years) and body mass index (BMI; <25 or ≥25 kg/m²). All 95% confidence intervals were calculated with 10,000 bootstrapping samples. In the image-level analysis, the AI model output was compared directly with the sonographer measurement in the same image, whereas in the study-level analysis, model outputs were compared with the clinical value reported in the final clinical echocardiography report. Data analysis was performed using Python (version 3.10.12). This study was carried out following the CONSORT-AI guideline^29^.

### Code and Data sharing Statements

The code for inference and fine-tuning, model weights, and demonstration video are available at https://github.com/echonet/measurements/. The patient data is not publicly available due to its potentially identifiable nature.

## Results

### Model Development and Validation Cohorts

A total of 186,712 images with segmentation information were used from 93,978 CSMC echocardiography studies from 56,855 patients as a derivation cohort. For temporal split validation, 66,955 images from 28,219 CSMC echocardiography studies (17,391 patients) were used. An additional 34,698 images from 28,777 KPNC echocardiography studies (20,186 patients) served as an external validation dataset. The patient characteristics in the CSMC derivation cohort were similar in patient age, female proportion, race, mean LVEF, and comorbidities across all datasets as temporal split dataset and external dataset (**Table 1**). The number of training and validation examples for each parameter is summarized in **Table 2**.

**Table 1.** Patient characteristics of the derivation, temporal split, and external validation cohorts.

|  | Derivation cohort<br>(CSMC, n = 56,855) | Temporal Split<br>Cohort (CSMC,<br>n = 17,391) | External<br>Validation<br>Cohort (KPNC,<br>n = 28,777) |
| --- | --- | --- | --- |
| N (patients) | 56855 | 17391 | 28777 |
| Age, years, median [IQR] | 68.0 [56.0–79.0] | 70.0 [60.0–79.0] | 70.00 [58.0–79.0] |
| Female sex, n (%) | 25936 (46.0%) | 7694 (44.2%) | 13978 (48.6%) |
| **Race/Ethnicity, n (%)** |  |  |  |
| White | 33532 (59.0%) | 9821 (56.5%) | 16145 (56.1%) |
| Black or African American | 7697 (13.5%) | 2512 (14.4%) | 2567 (8.9%) |
| Asian | 4084 (7.2%) | 1625 (9.3%) | 4338 (15.1%) |
| Hispanic/Latino | 7219 (12.7%) | 2187 (12.6%) | 4649 (16.2%) |
| Other/Unknown | 4323 (7.6%) | 1246 (7.2%) | 1078 (3.7%) |
| LVEF, %, median [IQR] | 62.0 [55.0–67.0] | 60.6 [52.8–65.4] | 60.0 [55.0–65.0] |
| **Comorbidities, n (%)** |  |  |  |
| Atrial Fibrillation | 4446 (7.8%) | 1317 (7.6%) | 8095 (28.1%) |
| Hypertension | 11098 (19.5%) | 4520 (26.0%) | 16582 (57.6%) |
| Diabetes mellitus | 6785 (11.9%) | 2599 (14.9%) | 9385 (32.6%) |
| Hyperlipidemia | 9712 (17.1%) | 5277 (30.3%) | 17149 (59.6%) |
| Coronary Artery Disease | 4368 (7.7%) | 1821 (10.5%) | 10583 (36.8%) |
| Heart Failure | 4477 (7.9%) | 1490 (8.6%) | 8249 (28.7%) |
| Stroke | 2083 (3.7%) | 464 (2.7%) | 2875 (10.0%) |

Data are presented as median [interquartile range] for continuous variables and as n (%) for categorical variables. LVEF = Left Ventricular Ejection Fraction.

**Table 2.** Dataset Characteristics.

| Measurement | Number of Images | Number of Studies | Number of Patients | Sonographer Measurement |
| --- | --- | --- | --- | --- |
| Derivation Cohort<br>(Patient n = 56855) |  |  |  |  |
| MV-VTI | 19,277 | 16,295 | 11,181 | 41.4 ± 15.3 cm |
| AV-VTI | 45,768 | 32,194 | 21,875 | 48.2 ± 24.4 cm |
| LVOT-VTI | 70,993 | 65,758 | 44,145 | 20.0 ± 7.4 cm |
| RVOT-VTI | 14,381 | 12,977 | 11,120 | 14.3 ± 4.5 cm |
| PV-VTI | 2,897 | 2,761 | 2,473 | 24.2 ± 13.7 cm |
| Left Atrium Area | 29,924 | 27,694 | 19,851 | 16.9 ± 8.9 cm<sup>2</sup> |
| Right Atrium Area | 12,621 | 12,601 | 10,088 | 13.8 ± 6.6 cm<sup>2</sup> |
| Temporal Split Cohort<br>(Patient n = 17391) |  |  |  |  |
| MV-VTI | 3,571 | 2,316 | 1,662 | 36.2 ± 16.6 cm |
| AV-VTI | 586 | 556 | 438 | 26.6 ± 14.7 cm |
| LVOT-VTI | 17,265 | 16,842 | 11,020 | 19.9 ± 6.1 cm |
| RVOT-VTI | 5,489 | 4,554 | 3,369 | 12.7 ± 4.6 cm |
| PV-VTI | 476 | 453 | 372 | 17.6 ± 6.4 cm |
| Left Atrium Area | 21,714 | 15,087 | 10,606 | 20.0 ± 6.5 cm<sup>2</sup> |
| Right Atrium Area | 6,744 | 5,947 | 4,250 | 16.4 ± 5.9 cm<sup>2</sup> |
| External Validation Cohort<br>( Patient n = 20,186) |  |  |  |  |
| MV-VTI | 1,386 | 1,266 | 1,223 | 38.8 ± 16.2 cm |
| AV-VTI | 6,677 | 5,463 | 4,961 | 45.3 ± 22.9 cm |
| LVOT-VTI | 2,820 | 2,666 | 2,542 | 24.5 ± 6.9 cm |
| RVOT-VTI | 126 | 126 | 124 | 19.4 ± 4.8 cm |
| PV-VTI | 542 | 527 | 522 | 23.6 ± 9.1 cm |
| Left Atrium Area | 21,069 | 19,635 | 15,338 | 20.2 ± 6.3 cm² |
| Right Atrium Area | 2,078 | 2,077 | 2,025 | 18.0 ± 7.2 cm² |

### Assessment of EchoNet-Segmentation Performance

In the CSMC held-out test dataset, the EchoNet-Segmentation models demonstrated strong agreement between automated deep learning measurements and sonographer measurements. For VTI measurements, R² value by EchoNet-Segmentation ranged from 0.817 to 0.882 and MAE ranged from 1.13 to 3.80 cm (**Figure 3**). Similarly, atrial area measurements achieved R² values between 0.675 and 0.747, with MAE between 2.48 and 2.52 cm² (**Figure 3**). Detailed information including comparison between mean values of sonographer measurements and deep learning-based measurements, R², and MAE in the held-out dataset are described in **Table 3** and the detailed metrics of YOLO VTI envelope detection model are summarized in **Supplemental Table 1**. Subgroup analyses stratified by atrial fibrillation status and body mass index demonstrated robust model performance across all subgroups for both VTI models and atrial area models (**Supplemental Table 2** and **Supplemental Table 3**). A demonstration video of frame-by-frame simultaneous RA and LA inference applied to an apical four-chamber view is shown in **Supplemental Video 1**.

**Figure 3.**
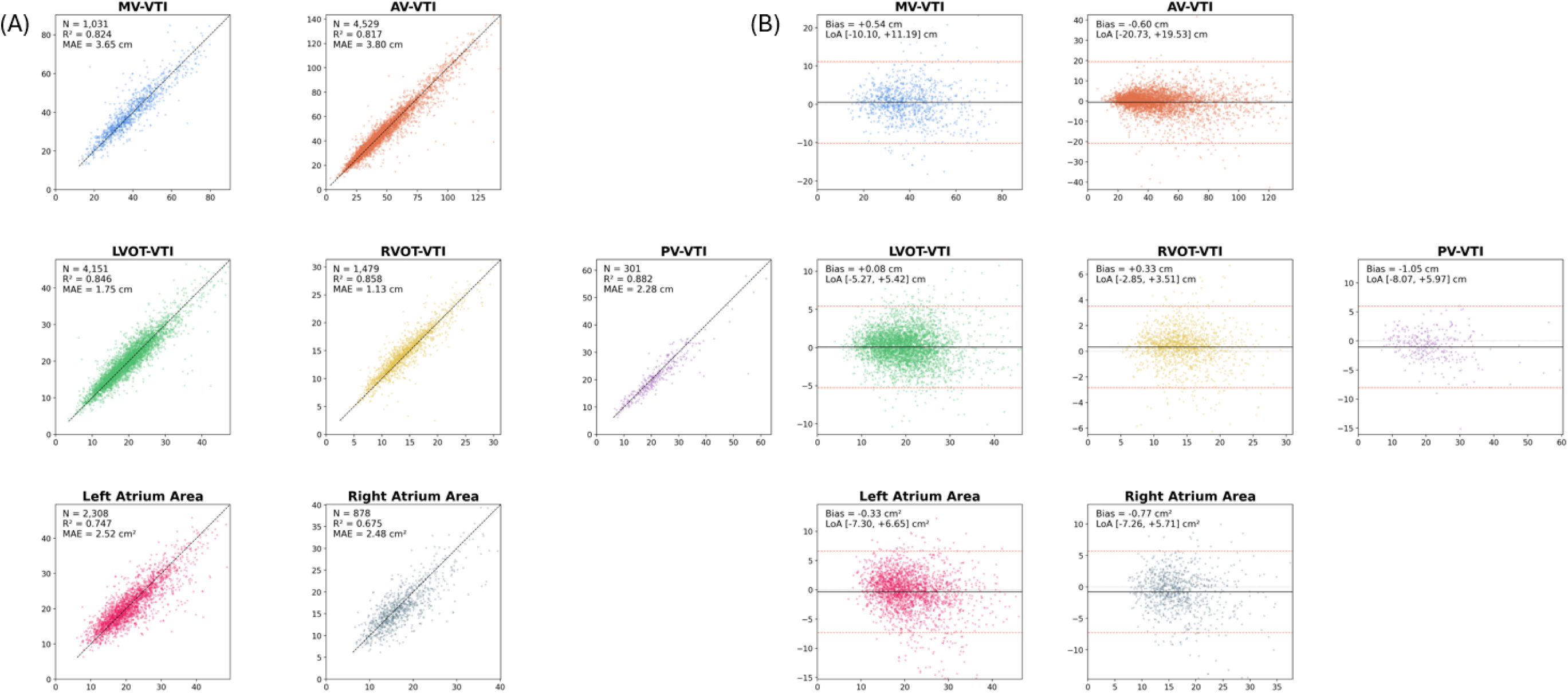
Performance of EchoNet-Segmentation on the CSMC held-out test dataset (Image-level analysis). For each parameter, the left panel (A) shows a scatter plot comparing automated deep learning measurements (y-axis) with sonographer reference measurements (x-axis). The right pan shows the corresponding Bland-Altman plot, in which the difference between the two measurements (y-axis) is plotted against their mean (x-axis); the solid line indicates the mean bias a dashed lines indicate the 95% limits of agreement. Detailed performance metrics are provided in **Table 3**.

**Table 3.** EchoNet-Segmentation Performance in Internal and External Test Datasets.

| Measurement | Number of Images | Sonographer value | AI value | Mean Absolute Error | R<sup>2</sup> | MAE 10th tile | MAE 50th tile | MAE 90th tile |
| --- | --- | --- | --- | --- | --- | --- | --- | --- |
| Held-out test in Derivation cohort (Image-level analysis) |  |  |  |  |  |  |  |  |
| MV-VTI | 1031 | 39.10 ± 12.59 cm | 39.64 ± 12.67 cm | 3.65 [3.41–3.90] | 0.824<br>[0.787–0.856] | 0.47 cm<br>[0.39–0.56] | 2.53 cm<br>[2.31–2.85] | 7.31 cm<br>[6.93–7.83] |
| AV-VTI | 4529 | 47.36 ± 24.03 cm | 46.76 ± 21.44 cm | 3.80 [3.56–4.10] | 0.817<br>[0.686–0.926] | 0.40 cm<br>[0.37–0.44] | 2.32 cm<br>[2.24–2.41] | 7.77 cm<br>[7.35–8.02] |
| LVOT-VTI | 4151 | 20.00 ± 6.94 cm | 20.08 ± 6.48 cm | 1.75 [1.69–1.81] | 0.846<br>[0.818–0.871] | 0.23 cm<br>[0.21–0.25] | 1.27 cm<br>[1.22–1.31] | 3.70 cm<br>[3.54–3.83] |
| RVOT-VTI | 1479 | 14.27 ± 4.26 cm | 14.60 ± 4.18 cm | 1.13 [1.07–1.20] | 0.858<br>[0.823–0.884] | 0.17 cm<br>[0.14–0.19] | 0.78 cm<br>[0.74–0.83] | 2.43 cm<br>[2.30–2.64] |
| PV-VTI | 301 | 22.64 ± 10.31 cm | 21.59 ± 9.15 cm | 2.28 [1.96–2.65] | 0.882<br>[0.791–0.937] | 0.30 cm<br>[0.22–0.43] | 1.72 cm<br>[1.43–1.91] | 4.69 cm<br>[3.72–5.35] |
| Left Atrium Area | 2308 | 21.33 ± 7.07 cm<sup>2</sup> | 21.01 ± 6.26 cm<sup>2</sup> | 2.52 [2.41–2.62] | 0.747<br>[0.718–0.775] | 0.32 cm<sup>2</sup><br>[0.29–0.37] | 1.79 cm<sup>2</sup><br>[1.70–1.89] | 5.42 cm<sup>2</sup><br>[5.13–5.77] |
| Right Atrium Area | 878 | 17.29 ± 5.71 cm<sup>2</sup> | 16.52 ± 5.29 cm<sup>2</sup> | 2.48 [2.33–2.65] | 0.675<br>[0.621–0.724] | 0.30 cm<sup>2</sup><br>[0.26–0.37] | 1.80 cm<sup>2</sup><br>[1.66–1.97] | 5.30 cm<sup>2</sup><br>[4.85–5.81] |
| Temporal Split Cohort (Study-level analysis) |  |  |  |  |  |  |  |  |
| MV-VTI | 3571 | 36.20 ± 16.62 cm | 37.76 ± 18.11 cm | 5.34 [4.99–5.66] | 0.616<br>[0.535–0.697] | 0.53 cm<br>[0.48–0.60] | 2.94 cm<br>[2.82–3.07] | 10.50 cm<br>[9.97–11.19] |
| AV-VTI | 586 | 26.56 ± 14.68 cm | 26.74 ± 13.18 cm | 3.43 [2.97–4.01] | 0.757<br>[0.630–0.868] | 0.37 cm<br>[0.30–0.43] | 1.87 cm<br>[1.71–2.11] | 6.59 cm<br>[5.43–8.10] |
| LVOT-VTI | 17265 | 19.90 ± 6.12 cm | 19.82 ± 5.93 cm | 1.60 [1.58–1.63] | 0.866<br>[0.853–0.875] | 0.21 cm<br>[0.20–0.22] | 1.20 cm<br>[1.18–1.22] | 3.43 cm<br>[3.36–3.49] |
| RVOT-VTI | 5489 | 12.68 ± 4.64 cm | 12.75 ± 4.91 cm | 1.72 [1.66–1.80] | 0.606<br>[0.537–0.661] | 0.16 cm<br>[0.14–0.17] | 1.00 cm<br>[0.96–1.05] | 3.83 cm<br>[3.64–3.95] |
| PV-VTI | 476 | 17.56 ± 6.42 cm | 16.72 ± 5.32 cm | 2.20 [1.98–2.44] | 0.731<br>[0.674–0.787] | 0.29 cm<br>[0.24–0.36] | 1.50 cm<br>[1.33–1.65] | 4.59 cm<br>[4.12–5.18] |
| Left Atrium Area | 21714 | 20.01 ± 6.54 cm<sup>2</sup> | 21.17 ± 6.15 cm<sup>2</sup> | 2.83 [2.80–2.86] | 0.705<br>[0.695–0.715] | 0.40 cm<sup>2</sup><br>[0.38–0.41] | 2.19 cm<sup>2</sup><br>[2.15–2.22] | 6.08 cm<sup>2</sup><br>[5.99–6.16] |
| Right Atrium Area | 6744 | 16.38 ± 5.95 cm<sup>2</sup> | 15.11 ± 5.19 cm<sup>2</sup> | 2.55 [2.50–2.61] | 0.694<br>[0.675–0.710] | 0.32 cm<sup>2</sup><br>[0.30–0.35] | 1.90 cm<sup>2</sup><br>[1.84–1.96] | 5.45 cm<sup>2</sup><br>[5.30–5.67] |
| External Validation Cohort<br>(Image-level analysis) |  |  |  |  |  |  |  |  |
| MV-VTI | 1386 | 38.79 ± 16.17 cm | 35.85 ± 13.99 cm | 5.91<br>[5.46–6.37] | 0.616<br>[0.565–0.668] | 0.52 cm<br>[0.46–0.59] | 3.18 cm<br>[2.99–3.51] | 12.55 cm<br>[10.99–14.97] |
| AV-VTI | 6677 | 45.26 ± 22.85 cm | 42.54 ± 21.55 cm | 4.47<br>[4.30–4.66] | 0.859<br>[0.841–0.878] | 0.42 cm<br>[0.39–0.44] | 2.36 cm<br>[2.29–2.43] | 8.89 cm<br>[8.47–9.40] |
| LVOT-VTI | 2820 | 24.46 ± 6.92 cm | 23.11 ± 6.34 cm | 2.56<br>[2.43–2.70] | 0.623<br>[0.571–0.670] | 0.26 cm<br>[0.24–0.30] | 1.55 cm<br>[1.47–1.62] | 5.12 cm<br>[4.86–5.34] |
| RVOT-VTI | 126 | 19.39 ± 4.77 cm | 18.82 ± 4.26 cm | 1.35<br>[1.09–1.63] | 0.824<br>[0.720–0.894] | 0.18 cm<br>[0.10–0.24] | 0.80 cm<br>[0.64–1.14] | 3.04 cm<br>[2.57–3.49] |
| PV-VTI | 542 | 23.56 ± 9.08 cm | 21.67 ± 7.31 cm | 3.52<br>[3.11–3.96] | 0.575<br>[0.442–0.694] | 0.37 cm<br>[0.27–0.51] | 2.20 cm<br>[2.02–2.45] | 6.28 cm<br>[5.61–7.96] |
| Left Atrium Area | 21069 | 20.22 ± 6.25 cm<sup>2</sup> | 21.28 ± 6.27 cm<sup>2</sup> | 2.19<br>[2.16–2.22] | 0.803<br>[0.790–0.814] | 0.32 cm<sup>2</sup><br>[0.30–0.33] | 1.71 cm<sup>2</sup><br>[1.68–1.73] | 4.48 cm<sup>2</sup><br>[4.42–4.55] |
| Right Atrium Area | 2078 | 18.04 ± 7.16 cm<sup>2</sup> | 18.73 ± 6.83<br>cm<sup>2</sup> | 1.79<br>[1.71–1.87] | 0.876<br>[0.851–0.895] | 0.24 cm<sup>2</sup><br>[0.20–0.28] | 1.31 cm<sup>2</sup><br>[1.27–1.38] | 3.63 cm<sup>2</sup><br>[3.41–3.87] |

On the temporal split dataset from CSMC, the model showed robust performance (e.g., R² of 0.866 and MAE of 1.60 cm for LVOT VTI, and R² of 0.705 and MAE of 2.83 cm² for LA area) (**Figure 4** and **Table 3**). In the external KPNC dataset, model performance was similarly robust (**Figure 4** and **Table 3**), with MAE ranging from 1.35 cm (RVOT-VTI) to 5.91 cm (MV-VTI) for VTI measurements and 1.79 cm² (RA area) to 2.19 cm² (LA area) for atrial area measurements. In the MIMIC-Echo dataset, we evaluated only AV VTI, LVOT VTI, LA volume, and RA length due to the availability of ground truth measurement value, demonstrating consistent model performance across different vendors (**Supplemental Table 4** and **Supplemental Figure 1**). Representative predicted images comparing sonographer annotations and deep learning annotations are shown in **Figure 5** for atrial area measurements and for VTI measurements.

**Figure 4:**
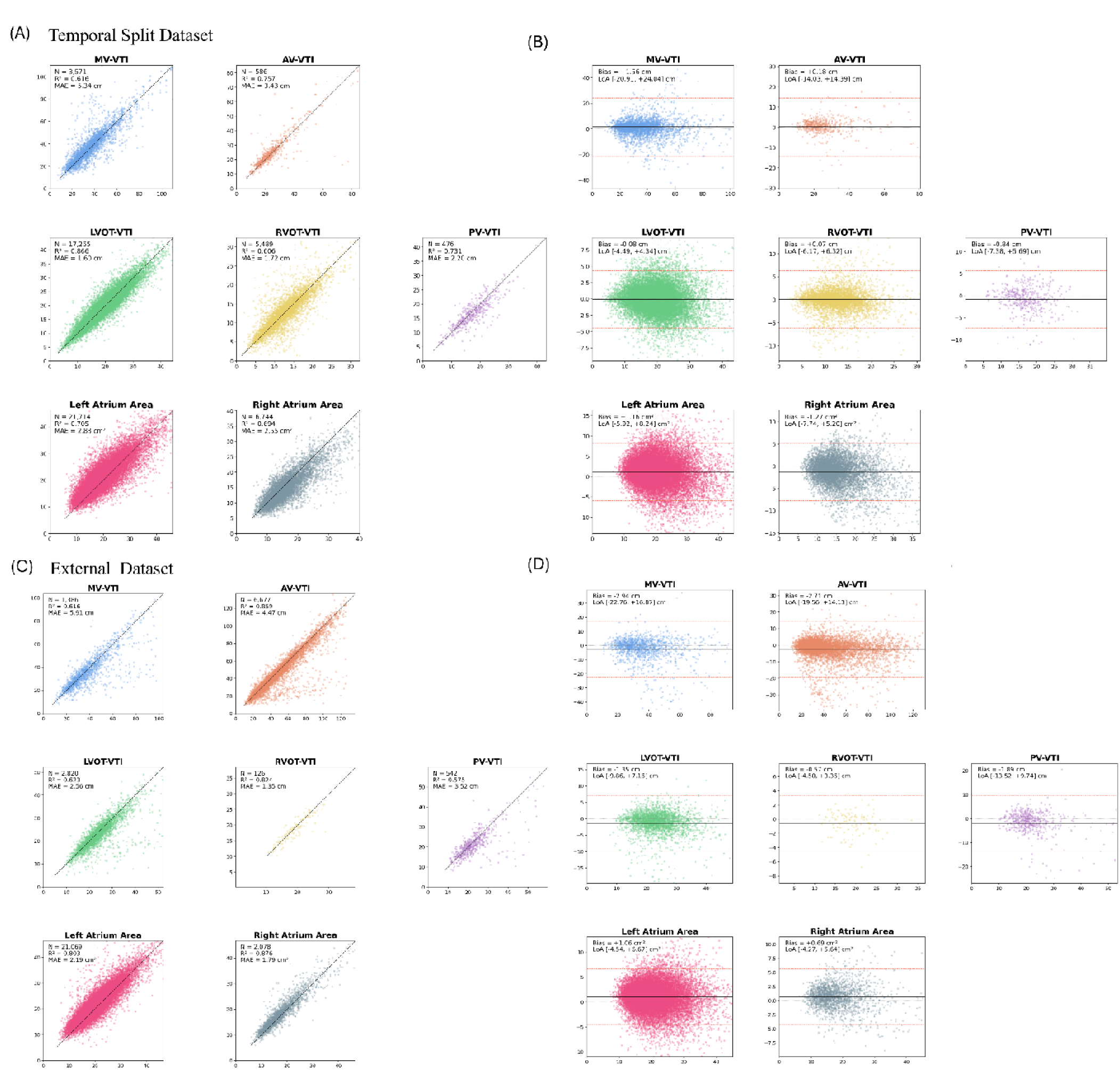
Performance of EchoNet-Segmentation on the CSMC temporal split dataset (Study-level analysis) and KPNC dataset (Image-level analysis) Performance of EchoNet-Segmentation on the temporal split cohort from Cedars-Sinai Medical Center (CSMC, A and B) and external validation dataset (Kaiser Permanente, C and D) in which echocardiography studies were analyzed using the end-to-end pipeline. For each parameter, the left panel (A and C) shows a scatter plot comparing automated deep learning measurements (y-axis) with clinical reference values (x-axis). The right panel (B and D) shows the corresponding Bland-Altman plot, in which the difference between the two measurements (y-axis) is plotted against their mean (x-axis); the solid line indicates the mean bias and the dashed lines indicate the 95% limits of agreement. Detailed performance metrics are provided in **Table 3**.

**Figure 5.**
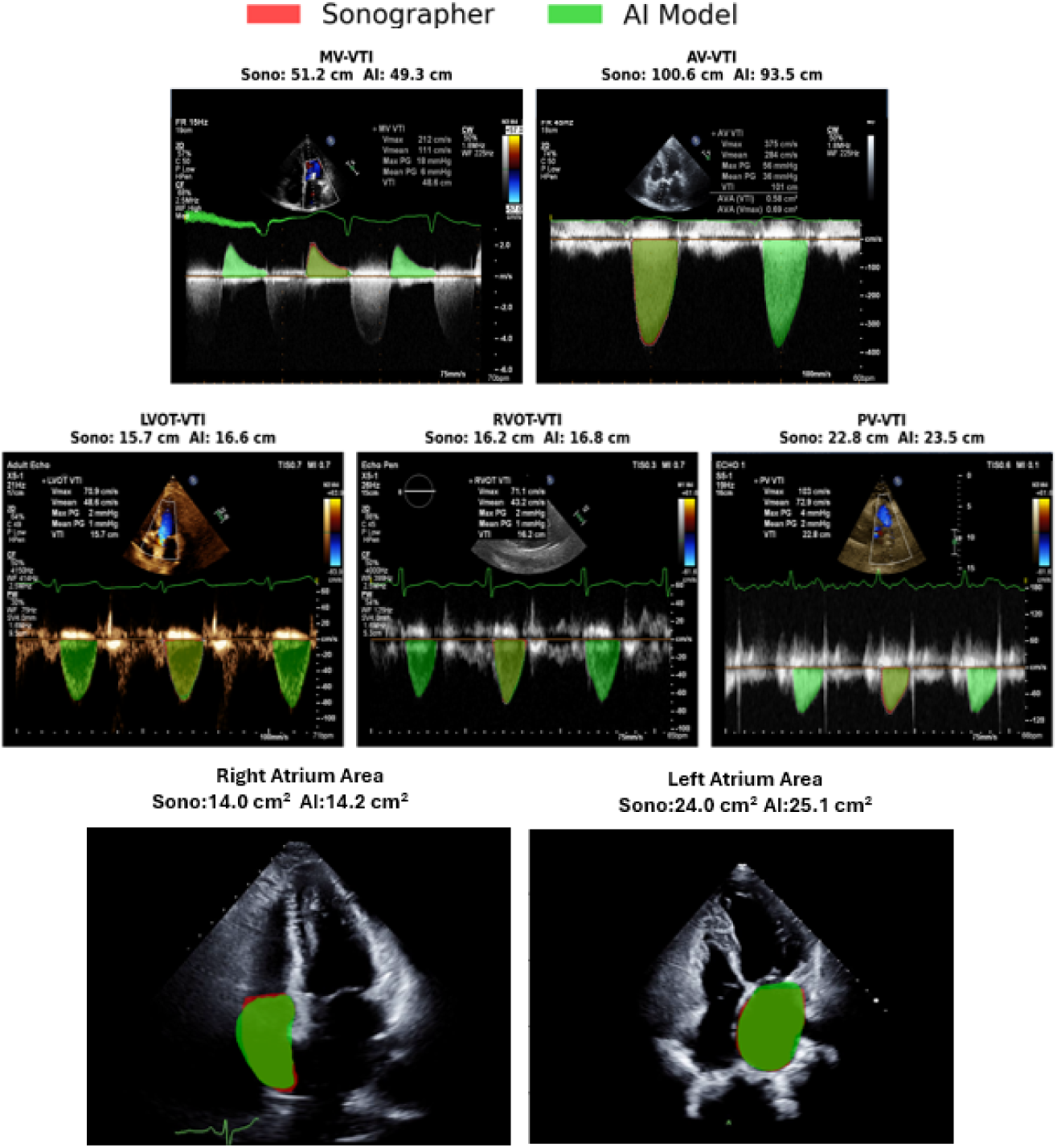
Representative images comparing sonographer and deep learning–based segmentations. Representative echocardiographic images illustrate the agreement between sonographer reference annotations and EchoNet-Segmentation outputs. For each parameter, the red mask shows the sonographer-annotated reference, and the green mask shows the corresponding AI based segmentation.

Model performance was robust and generalizable across a wide range of echocardiography formats and display configurations. We confirmed consistent automated measurements on screenshots from echocardiography machine monitors, images with side-by-side Doppler layouts, and images with various color patterns for Doppler spectral displays (**Supplemental Figure 2**). Demonstration videos of an interactive box-prompt-based VTI segmentation demo using EchoNet-Segmentation, are publicly released (**Supplemental Video 2**).

On the CSMC internal held-out test set, EchoNet-Segmentation outperformed MedSAM2 with both bounding-box and point prompts on R², MAE and Dice score, while zero-shot MedSAM2 (whole image, no prompt) failed to produce usable segmentations (**Table 4**, **Supplemental Figure 3**). We further validated the performance on a randomly sampled MIMIC-ECHO apical four-chamber subset (LA Area: n = 300, RA Area: n = 300) with manually traced ground-truth masks by board-certified cardiologists. On this external dataset, EchoNet-Segmentation outperformed MedSAM2 with bounding-box and point prompts across R², MAE and Dice score for both traced LA and RA area (**Supplemental Table 5**, **Supplemental Figures 4 and 5**). The bounding-box and point-prompt coordinates and the manual segmentation masks used for these comparisons in MIMIC-ECHO were publicly released (see **Code and Data Availability**).

**Table 4.**
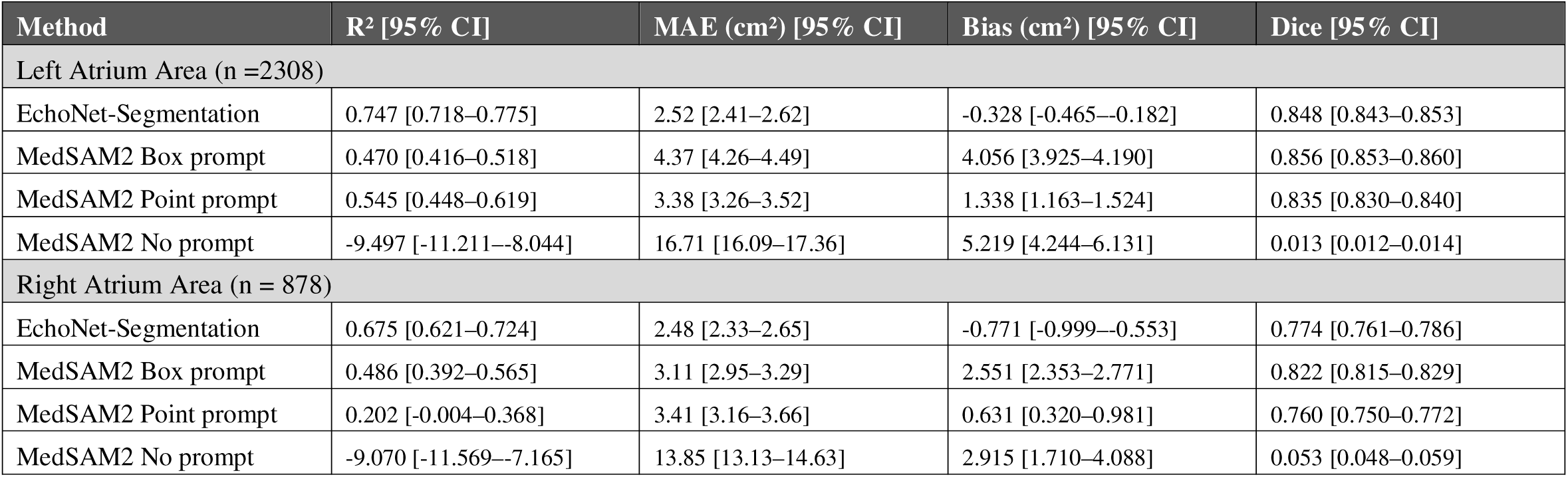
Comparison between MedSAM2 and EchoNet-Segmentation performance (held-out dataset).

## Discussion

In this study, we developed EchoNet-Segmentation, a deep learning model that automatically segments seven distinct echocardiographic parameters and maintains strong performance when evaluated on cohorts from different institutions, time periods, and public data. Trained on the largest real-world collection of sonographer-annotated segmentations to date, the model reached an accuracy comparable to that of expert sonographers. The code, model weights, and an interactive demo interface have been released to facilitate research and to support clinical deployment.

Comprehensive echocardiography measurement is required since clinical echocardiography involves the assessment of many parameters that holistically reflect patient status. Cardiovascular disease includes a wide spectrum such as left and right ventricular dysfunction, valvular and myocardial disease, and congenital heart disease. Despite this clinical breadth, existing open-source models have covered only a limited subset of measurements. While some deep learning models for VTI and atrial area segmentation are reported^17,19,30,31^, the code and model weights have largely remained unavailable, limiting reproducibility and downstream application. In the present study, we compared EchoNet-Segmentation with an open-source foundation model (MedSAM2) for medical image segmentation with fine-tuning using apical four-chamber segmentation. However, generating valid atrial area predictions with MedSAM2 requires manual prompt input (**Supplemental Figure 3**), which could be a structural limitation that precludes high-throughput and fully automated analysis. In contrast, EchoNet-Segmentation does not require any manual prompting and can be applied directly to large image cohorts, making it well suited for high-throughput, fully automated analysis. Furthermore, our task-specific EchoNet-Segmentation model outperformed this medical imaging foundation model on benchmarking against publicly available data.

Together with EchoNet-Measurements, EchoPrime and EchoNet-Dynamic^12,15^, the present work brings an automated suite of tools that produce a preliminary report and 26 publicly available task-specific an models to, enabling an end-to-end clinical workflow including image acquisition, view classification, and view-specific measurement. Clinically, this supports high-throughput automation of guideline-recommended assessments, including aortic stenosis severity grading by the continuity equation, heart failure with preserved ejection fraction diagnosis through left atrial volume assessment, shunt quantification, and evaluation of diastolic function.

This study has several strengths. First, to our knowledge it represents the largest and most comprehensive suite of task-specific echocardiographic segmentation models developed to date, leveraging real-world sonographer annotations from routine clinical practice. Second, generalizability was demonstrated across vendors and image formats, with robust performance maintained in subgroup analyses. Third, the model was directly benchmarked against an existing foundation model, enabling a transparent comparison with prior approaches.

There are some limitations in the present study. First, the development dataset was obtained from a single center, and although robust performance was confirmed on data from other vendors, the training data included single vendor machines. Second, the VTI models could not be compared with other AI-based measurement tools owing to limited public availability and the absence of an established benchmark dataset. Benchmark datasets and prospective randomized controlled trials, similar to prior work, will be required to further validate clinical utility.

## Conclusion

In this study, we developed and validated EchoNet-Segmentation, an open-source deep learning framework for the automated segmentation of a wide range of echocardiographic parameters. The model demonstrated robust performance across several echocardiographic parameters in both internal and external datasets.

## Contributions

YS and DO designed the study. YS, DC, and MV developed and trained the deep learning models. YS, HI, and MR curated and annotated the echocardiographic data. YS and BH performed the statistical analysis. YS, ACK, SC, and DO interpreted the results. YS drafted the manuscript, and all authors critically revised it for important intellectual content. DO supervised the study and obtained funding. All authors approved the final version of the manuscript.

## Supporting information

Supplemental Doc

## Acknowledgements

We appreciate Dr. Hajime Ichiryu for providing a side-by-side layout echocardiography Doppler image.

---

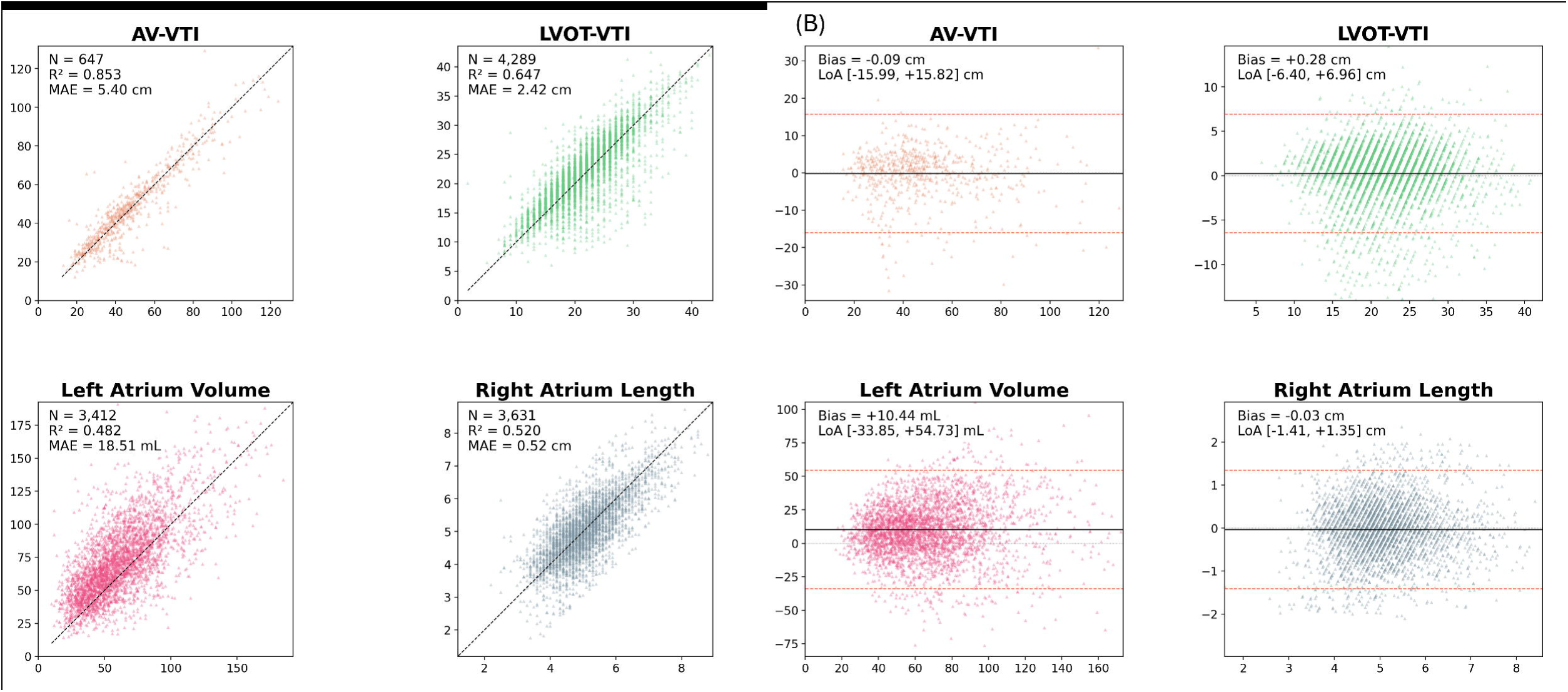

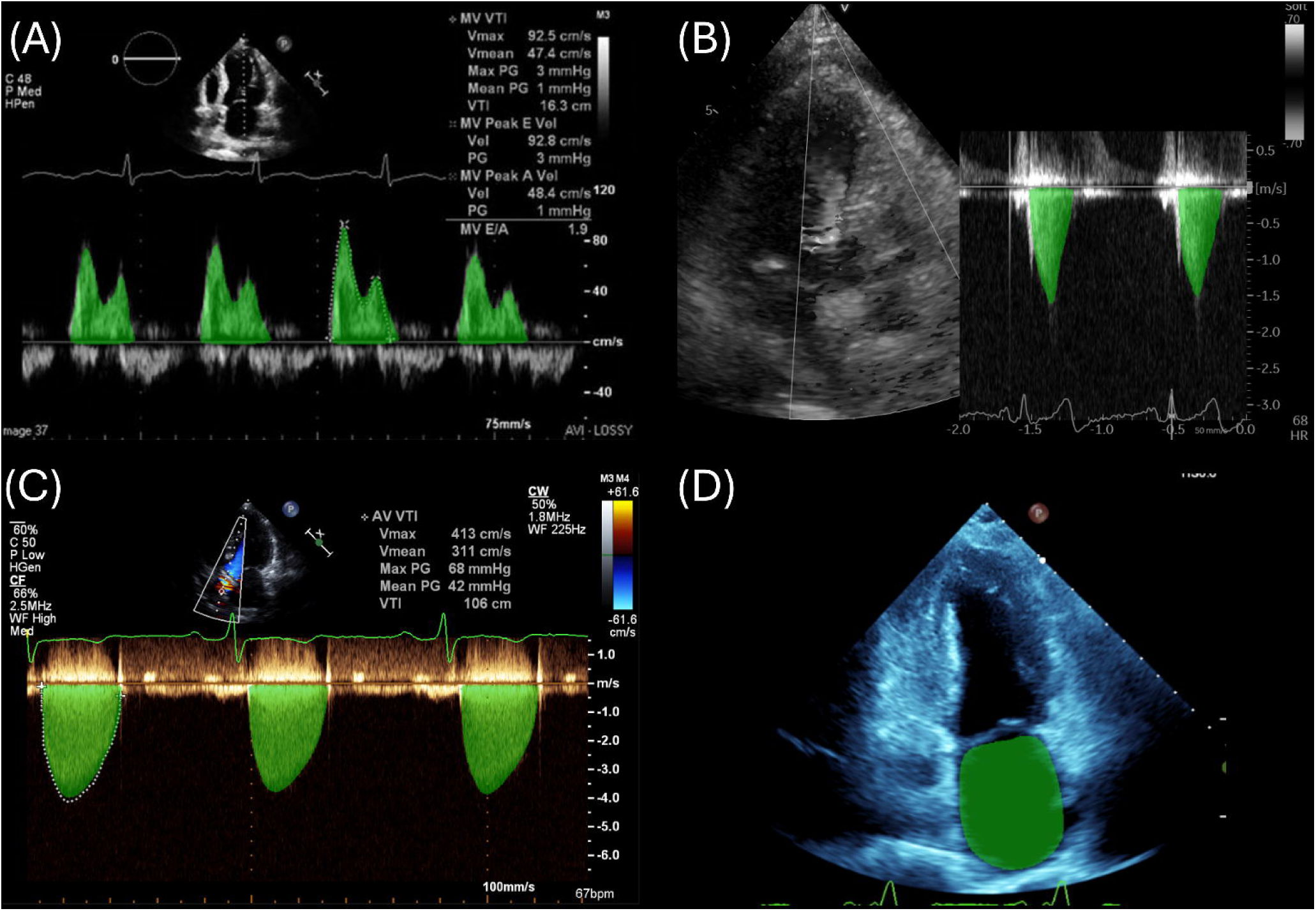

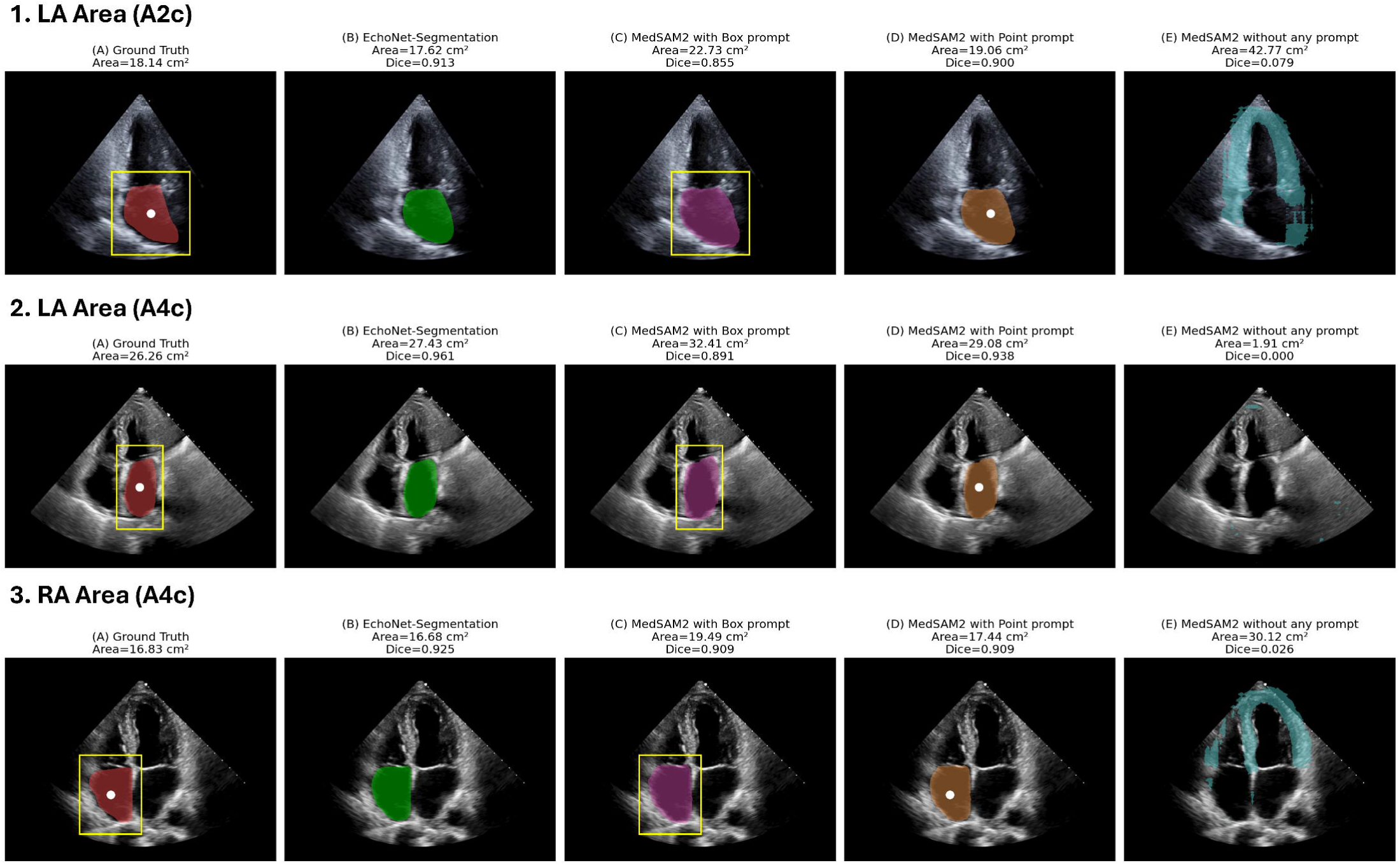

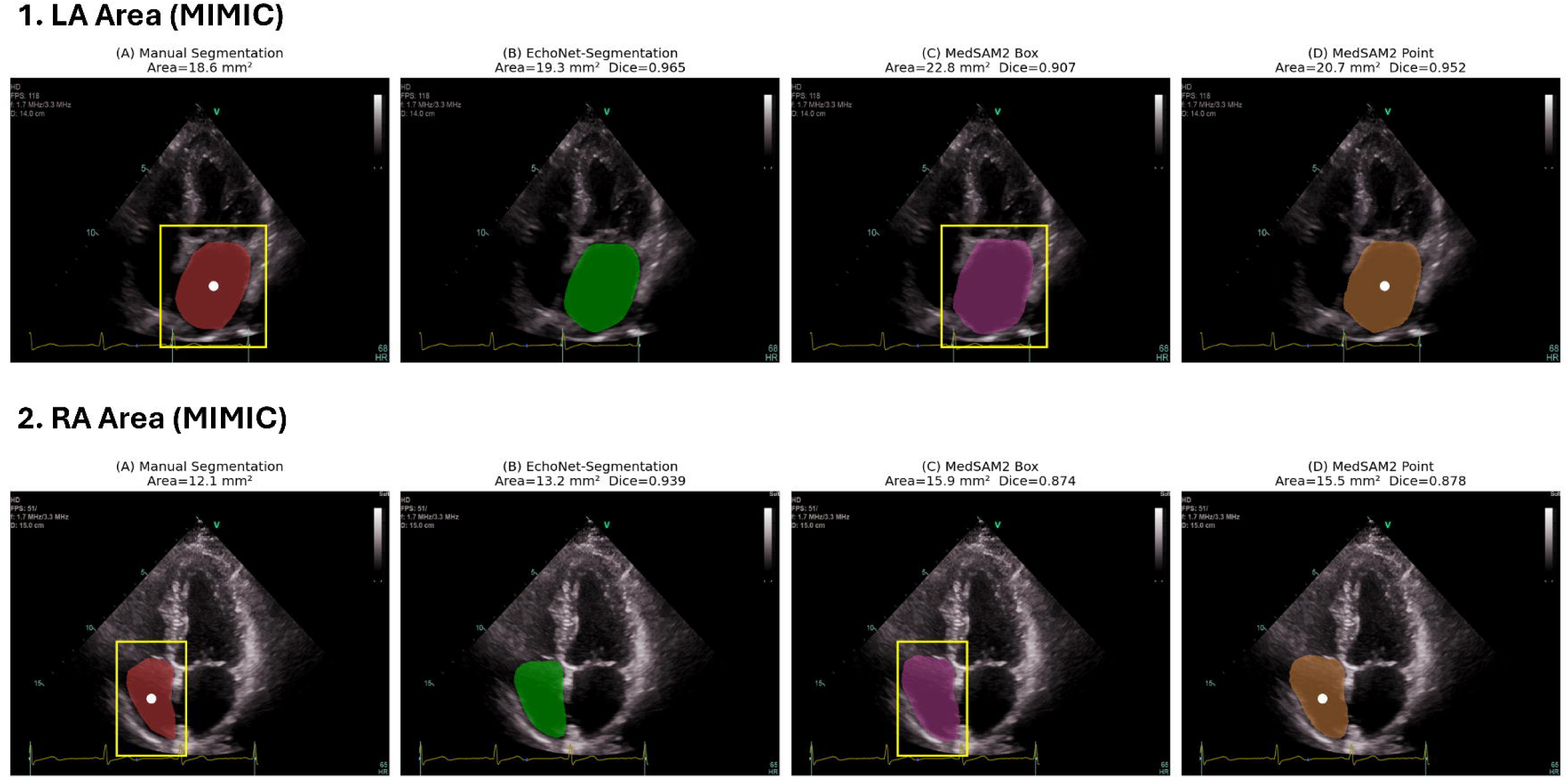

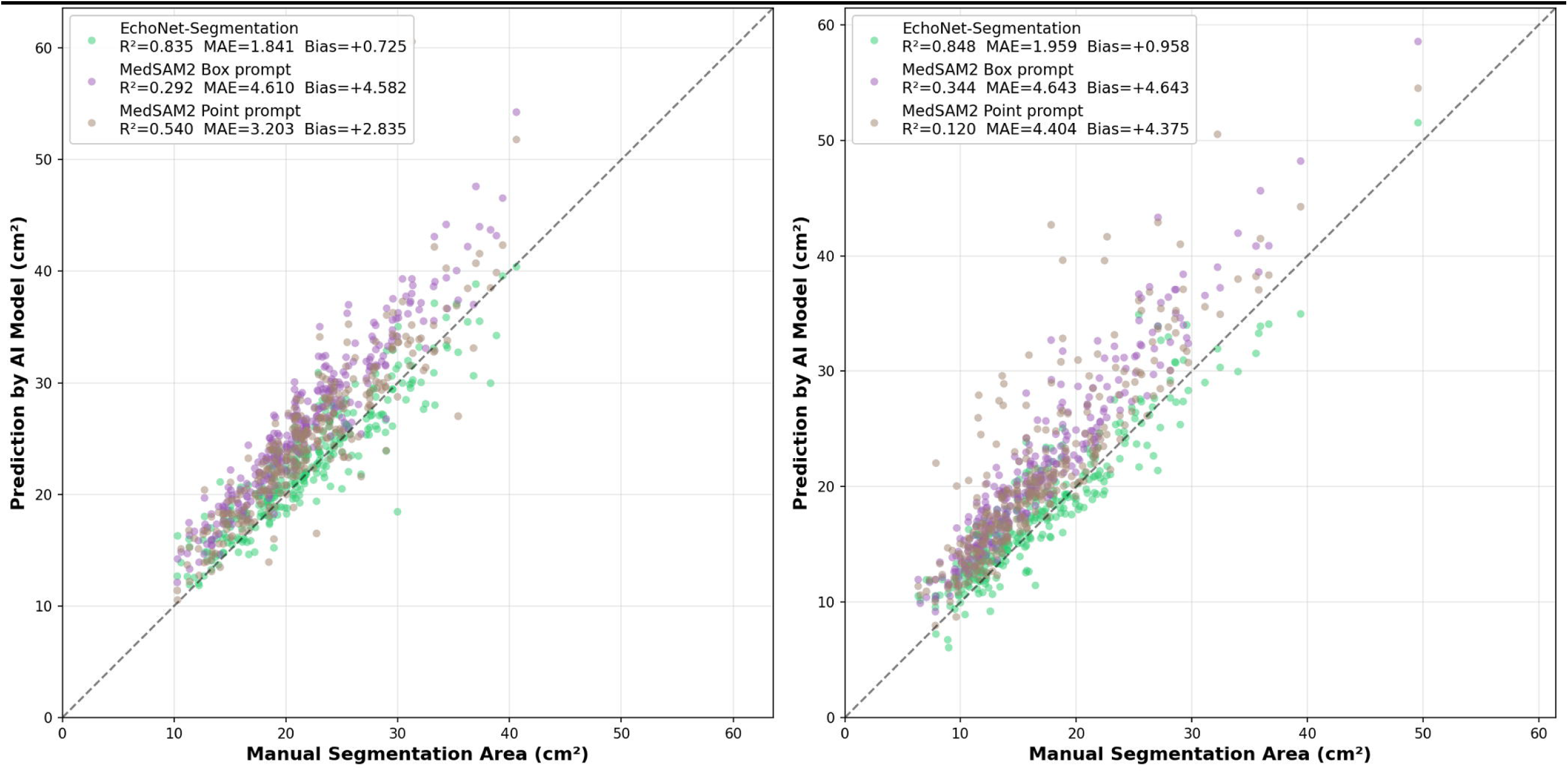

