## Supplemental Doc for "Multi-task artificial intelligence annotation of echocardiographic images: a retrospective multi-cohort study"

Authors and affiliations:

Yuki Sahashi MD, MSc, PhD^1^, David Choi BA^1^, Hirotaka Ieki MD, PhD^2,3^, Milos Vukadinovic BS^3^, Meenal Rawlani BS^1^, Bryan He PhD^4^, Alan C. Kwan MD^1^, Susan Cheng MD, MSc^1^, David Ouyang MD^1,5^

**Supplemental Table 1**: Object detection performance of the YOLO model for VTI envelope localization

| **Measurement** | **Number of images** | **mAP@50** | **mAP@50-95** | **Mean IoU** |
| --- | --- | --- | --- | --- |
| MV-VTI | 1031 | 0.955 | 0.884 | 0.867 |
| AV-VTI | 4529 | 0.970 | 0.918 | 0.872 |
| LVOT-VTI | 4151 | 0.966 | 0.893 | 0.878 |
| RVOT-VTI | 1479 | 0.946 | 0.855 | 0.843 |
| PV-VTI | 301 | 0.974 | 0.915 | 0.882 |

**Supplemental Table 2**. EchoNet-Segmentation performance stratified by atrial fibrillation status in the CSMC held-out test dataset.

| **Measurement** | **Subgroup** | **Number of Images** | **Sonographer value** | **AI value** | **Mean Absolute Error** | **R²** |
| --- | --- | --- | --- | --- | --- | --- |
| MV VTI | No AF | 963 | 39.09 ± 12.53 cm | 39.67 ± 12.66 cm | 3.73 [3.48–4.00] | 0.802 [0.757–0.844] |
|  | AF | 68 | 39.18 ± 13.46 cm | 39.21 ± 12.79 cm | 2.53 [1.97–3.12] | 0.933 [0.891–0.956] |
| AV VTI | No AF | 4327 | 47.46 ± 24.23 cm | 46.91 ± 21.62 cm | 3.79 [3.54–4.11] | 0.818 [0.683–0.929] |
|  | AF | 202 | 45.24 ± 19.02 cm | 43.70 ± 16.98 cm | 3.97 [3.03–5.34] | 0.758 [0.531–0.941] |
| LVOT VTI | No AF | 3979 | 20.12 ± 6.94 cm | 20.20 ± 6.48 cm | 1.74 [1.68–1.80] | 0.847 [0.816–0.875] |
|  | AF | 172 | 17.22 ± 6.38 cm | 17.29 ± 5.81 cm | 1.86 [1.52–2.27] | 0.771 [0.615–0.897] |
| RVOT VTI | No AF | 1406 | 14.35 ± 4.26 cm | 14.67 ± 4.15 cm | 1.13 [1.07–1.20] | 0.850 [0.813–0.878] |
|  | AF | 73 | 12.87 ± 4.06 cm | 13.29 ± 4.57 cm | 1.06 [0.73–1.45] | 0.796 [0.599–0.927] |
| PV VTI | No AF | 285 | 22.86 ± 10.43 cm | 21.82 ± 9.25 cm | 2.31 [1.97–2.69] | 0.868 [0.766–0.926] |
|  | AF | 16 | 18.75 ± 6.80 cm | 17.59 ± 5.62 cm | 1.92 [1.09–2.87] | 0.852 [0.644–0.941] |
| Left Atrium Area | No AF | 2215 | 21.10 ± 6.86 cm² | 20.78 ± 6.09 cm² | 2.50 [2.40–2.59] | 0.736 [0.705–0.765] |
|  | AF | 93 | 26.98 ± 9.26 cm² | 26.42 ± 7.55 cm² | 2.97 [2.33–3.72] | 0.752 [0.597–0.861] |
| Right Atrium Area | No AF | 838 | 17.10 ± 5.60 cm² | 16.35 ± 5.18 cm² | 2.45 [2.30–2.63] | 0.643 [0.573–0.700] |
|  | AF | 40 | 21.31 ± 6.41 cm² | 20.02 ± 6.24 cm² | 3.15 [2.29–4.09] | 0.551 [0.111–0.768] |

**Supplemental Table 3**. EchoNet-Segmentation performance stratified by body mass index in the CSMC held-out test dataset.

| **Measurement** | **Subgroup** | **Number of Images** | **Sonographer value** | **AI value** | **Mean Absolute Error** | **R²** |
| --- | --- | --- | --- | --- | --- | --- |
| MV VTI | BMI <25 | 362 | 38.26 ± 12.48 cm | 38.67 ± 12.14 cm | 3.83 [3.39–4.33] | 0.765 [0.664–0.845] |
|  | BMI ≥25 | 570 | 39.69 ± 12.48 cm | 40.27 ± 12.74 cm | 3.60 [3.32–3.89] | 0.835 [0.795–0.871] |
|  | BMI unknown | 99 | 38.73 ± 13.43 cm | 39.53 ± 13.88 cm | 3.31 [2.58–4.14] | 0.840 [0.703–0.929] |
| AV VTI | BMI <25 | 1694 | 46.25 ± 22.04 cm | 45.53 ± 20.76 cm | 3.71 [3.44–4.02] | 0.898 [0.855–0.935] |
|  | BMI ≥25 | 2429 | 47.97 ± 25.54 cm | 47.36 ± 21.84 cm | 3.86 [3.47–4.37] | 0.759 [0.586–0.933] |
|  | BMI unknown | 406 | 48.40 ± 22.38 cm | 48.33 ± 21.53 cm | 3.78 [3.41–4.22] | 0.936 [0.907–0.957] |
| LVOT VTI | BMI <25 | 1638 | 19.62 ± 6.42 cm | 19.65 ± 5.96 cm | 1.70 [1.60–1.80] | 0.824 [0.779–0.863] |
|  | BMI ≥25 | 2151 | 20.43 ± 7.37 cm | 20.47 ± 6.84 cm | 1.81 [1.72–1.91] | 0.851 [0.808–0.889] |
|  | BMI unknown | 362 | 19.17 ± 6.42 cm | 19.67 ± 6.40 cm | 1.60 [1.46–1.75] | 0.889 [0.852–0.917] |
| RVOT VTI | BMI <25 | 545 | 13.70 ± 3.98 cm | 14.04 ± 3.93 cm | 1.06 [0.97–1.17] | 0.845 [0.795–0.883] |
|  | BMI ≥25 | 801 | 14.61 ± 4.41 cm | 14.94 ± 4.30 cm | 1.18 [1.10–1.27] | 0.844 [0.786–0.882] |
|  | BMI unknown | 133 | 14.63 ± 4.21 cm | 14.93 ± 4.15 cm | 1.09 [0.92–1.27] | 0.870 [0.807–0.912] |
| PV VTI | BMI <25 | 111 | 21.24 ± 8.34 cm | 20.64 ± 7.74 cm | 2.19 [1.80–2.64] | 0.849 [0.724–0.923] |
|  | BMI ≥25 | 169 | 23.17 ± 11.17 cm | 21.90 ± 9.77 cm | 2.31 [1.86–2.85] | 0.866 [0.692–0.943] |
|  | BMI unknown | 21 | 25.77 ± 11.31 cm | 24.14 ± 10.16 cm | 2.63 [1.93–3.36] | 0.926 [0.786–0.960] |
| Left Atrium Area | BMI <25 | 886 | 20.19 ± 6.72 cm² | 20.15 ± 6.12 cm² | 2.32 [2.16–2.47] | 0.755 [0.706–0.803] |
|  | BMI ≥25 | 1180 | 22.58 ± 7.26 cm² | 21.99 ± 6.38 cm² | 2.62 [2.48–2.78] | 0.741 [0.698–0.779] |
|  | BMI unknown | 242 | 19.47 ± 6.26 cm² | 19.36 ± 5.26 cm² | 2.71 [2.37–3.05] | 0.617 [0.525–0.702] |
| Right Atrium Area | BMI <25 | 312 | 16.33 ± 5.27 cm² | 15.88 ± 4.91 cm² | 2.25 [2.00–2.48] | 0.649 [0.544–0.734] |
|  | BMI ≥25 | 472 | 18.01 ± 5.75 cm² | 17.02 ± 5.38 cm² | 2.69 [2.48–2.92] | 0.597 [0.496–0.679] |
|  | BMI unknown | 94 | 16.86 ± 6.37 cm² | 16.11 ± 5.76 cm² | 2.21 [1.83–2.62] | 0.787 [0.670–0.862] |

**Supplemental Table 4**. EchoNet-Segmentation Performance in MIMIC dataset.

| **Measurement** | **Number of Images** | **Sonographer value** | **AI value** | **Mean Absolute Error** | **R²** | **MAE 10th** | **MAE 50th** | **MAE 90th** |
| --- | --- | --- | --- | --- | --- | --- | --- | --- |
| AV VTI | 647 | 48.48 ± 20.80 cm | 48.39 ± 20.73 cm | 5.40 [4.96–5.89] | 0.853 [0.818–0.884] | 0.62 cm [0.45–0.85] | 3.57 cm [3.23–3.95] | 12.33 cm [10.87–13.54] |
| LVOT VTI | 4289 | 21.49 ± 5.45 cm | 21.77 ± 5.44 cm | 2.42 [2.34–2.49] | 0.647 [0.621–0.672] | 0.37 cm [0.33–0.40] | 1.76 cm [1.70–1.82] | 5.10 cm [4.88–5.40] |
| Left Atrium Volume* | 3412 | 62.22 ± 27.65 mL | 72.66 ± 29.93 mL | 18.51 [17.95–19.05] | 0.482 [0.447–0.516] | 2.63 mL [2.32–2.92] | 14.65 mL [14.08–15.16] | 39.51 mL [38.27–40.81] |
| Right Atrium Length* | 3631 | 5.10 ± 0.91 cm | 5.07 ± 0.97 cm | 0.52 [0.51–0.54] | 0.520 [0.486–0.554] | 0.07 cm [0.06–0.07] | 0.40 cm [0.38–0.41] | 1.15 cm [1.11–1.19] |

*Study-level evaluation was performed for LA volume, and RA length instead of atrial area, as LA area and RA area are not reported in the MIMIC clinical record.

**Supplemental Table 5**. Comparison between MedSAM2 and EchoNet-Segmentation performance (MIMIC).

| **Method** | **R² [95% CI]** | **MAE (cm²) [95% CI]** | **Bias (cm²) [95% CI]** | **Dice [95% CI]** |
| --- | --- | --- | --- | --- |
| Left Atrium Area (n = 300) | | | | |
| EchoNet-Segmentation | 0.835 [0.784–0.873] | 1.841 [1.671–2.010] | 0.725 [0.480–0.987] | 0.904 [0.892–0.916] |
| MedSAM2 Box prompt | 0.292 [0.142–0.408] | 4.610 [4.389–4.847] | 4.582 [4.348–4.827] | 0.896 [0.893–0.900] |
| MedSAM2 Point prompt | 0.540 [0.327–0.672] | 3.203 [2.947–3.483] | 2.835 [2.531–3.176] | 0.906 [0.901–0.910] |
| Right Atrium Area (n =300) | | | | |
| EchoNet-Segmentation | 0.848 [0.793–0.883] | 1.959 [1.798–2.148] | 0.958 [0.718–1.233] | 0.890 [0.875–0.902] |
| MedSAM2 Box prompt | 0.344 [0.157–0.491] | 4.643 [4.384–4.904] | 4.643 [4.384–4.904] | 0.871 [0.866–0.877] |
| MedSAM2 Point prompt | 0.120 [-0.254–0.386] | 4.404 [3.939–4.872] | 4.375 [3.903–4.839] | 0.859 [0.849–0.870] |

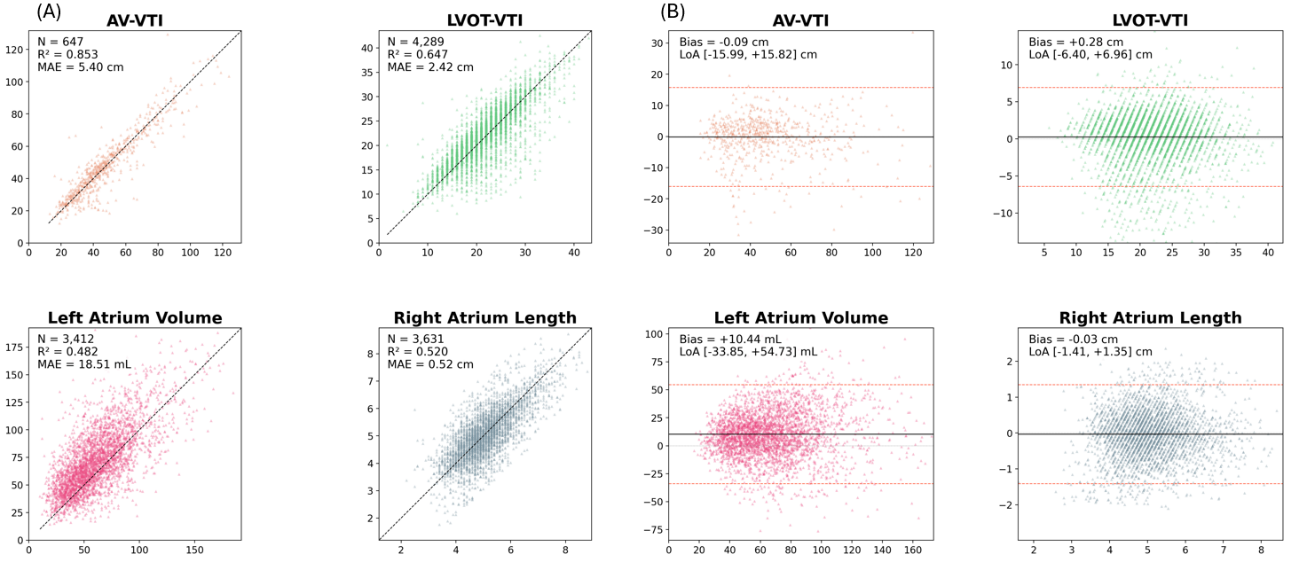
**Supplemental Figure 1**: Performance of EchoNet-Segmentation on the MIMIC-IV-ECHO validation cohort (Study-level analysis).

Performance of EchoNet-Segmentation on the MIMIC-IV-ECHO cohort, in which echocardiography studies were analyzed using the end-to-end pipeline. Because per-image sonographer annotations are not available in MIMIC, only volume and length measurements that can be derived from segmented structures were evaluated. For each parameter, the left panel (A) shows a scatter plot comparing automated deep learning measurements (y-axis) with the clinical reference values (x-axis). The right panel (B) shows the corresponding Bland-Altman plot. Detailed performance metrics are provided in **Supplemental Table 4**.

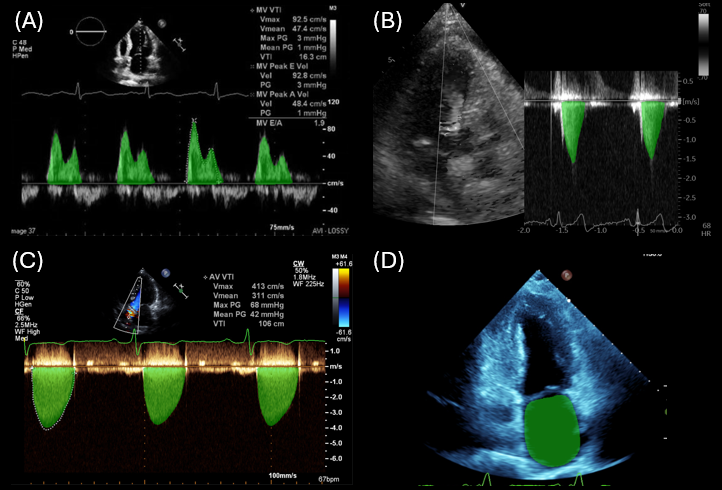
**Supplemental Figure 2**: Representative Images of EchoNet-Segmentation Inference across diverse image variations.

Model performance across non-standard image formats. (A) A screenshot captured from a website. (B) A Doppler tracing (LVOT-VTI) shown in a side-by-side layout. (C) An aortic valve VTI image rendered with a Chroma map color variation. (D) An apical two-chamber (A2C) LA area image, also rendered with Chroma map.

**Supplemental Figure 3**: Representative Comparison Inferenced Images between EchoNet-Segmentation and MedSAM2.

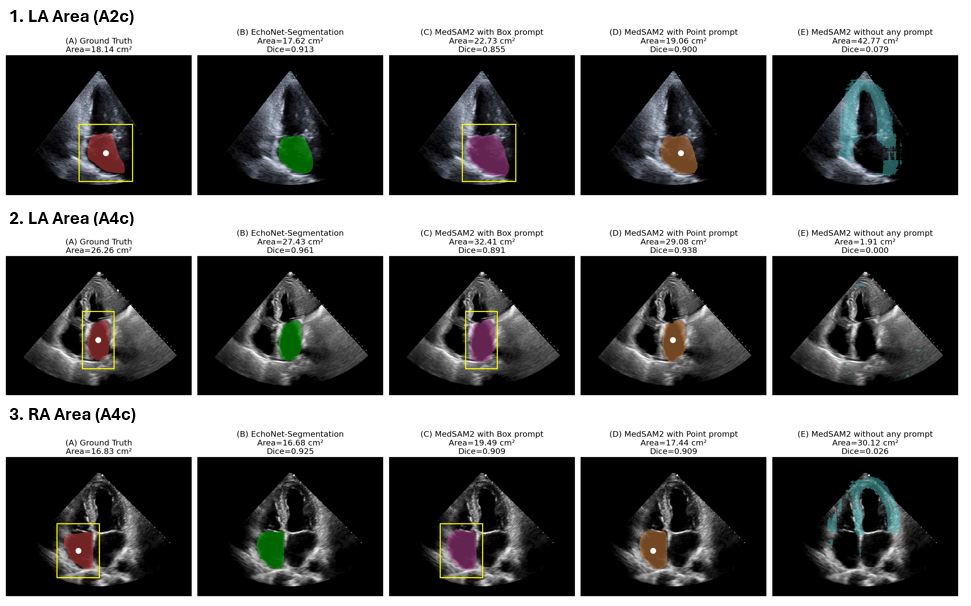

1. LA area in the apical two-chamber view (A2C), 2. LA area in the apical four-chamber view (A4C), and 3. RA area in the A4C view. For each case, the following are shown: (A) Ground truth segmentation by an expert sonographer, together with the bounding box (yellow) and point prompt (white dot); (B) Inference by EchoNet-Segmentation, EchoNet-Segmentation predicts segmentations directly from the entire image without any user-provided prompt. (C) Inference by MedSAM2 using the bounding-box prompt; (D) Inference by MedSAM2 using the point prompt; and (E) Inference by MedSAM2 without any prompts. All datasets are from held-out test dataset (CSMC).

**Supplemental Figure 4**: Representative Comparison Inferenced Images between EchoNet-Segmentation and MedSAM2 (MIMIC Dataset).

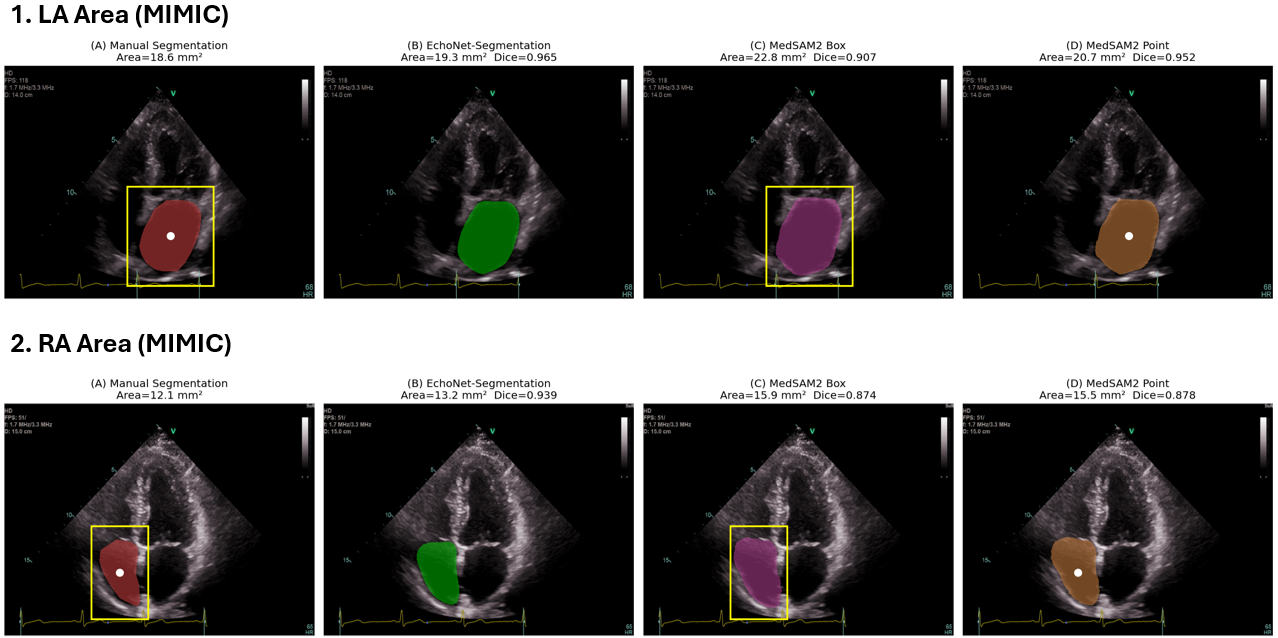

1. LA area in the apical two-chamber view (A2C), 2. LA area in the apical four-chamber view (A4C), and 3. RA area in the A4C view. For each case, the following are shown: (A) Ground truth segmentation by an expert sonographer, together with the bounding box (yellow) and point prompt (white dot); (B) Inference by EchoNet-Segmentation, EchoNet-Segmentation predicts segmentations directly from the entire image without any user-provided prompt. (C) Inference by MedSAM2 using a bounding-box prompt (yellow) and (D) Inference by MedSAM2 using a point prompt (white). All datasets are from MIMIC Dataset. Segmentation mask information, box prompt coordinates, and point prompt coordinates are released publicly.

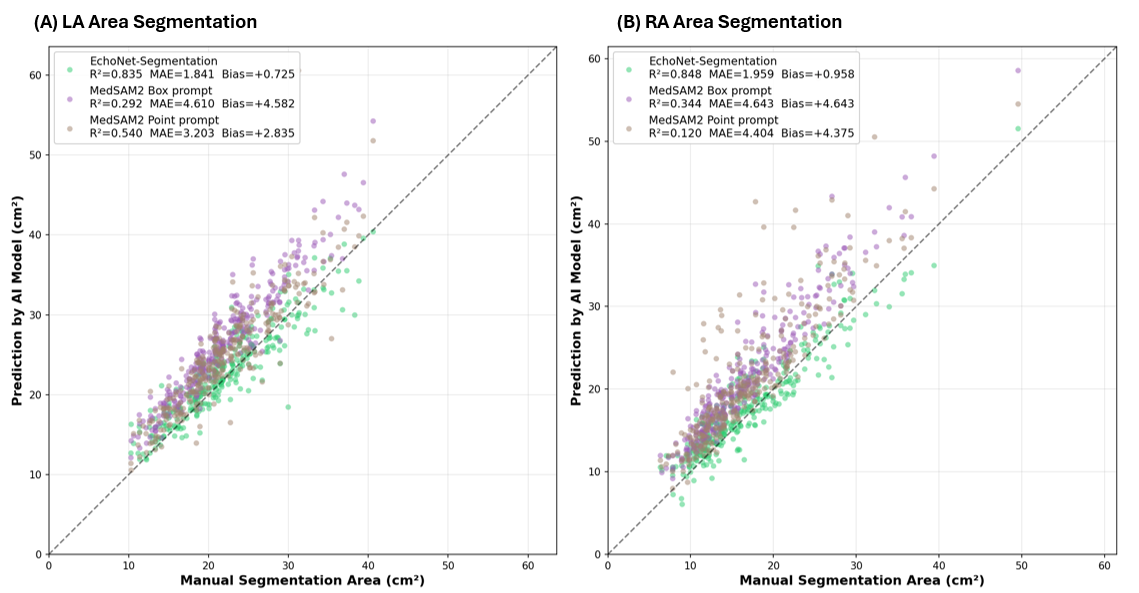
**Supplemental Figure 5**: Performance of EchoNet-Segmentation and MedSAM2 on the MIMIC-IV-ECHO cohort (Image-level analysis).

For segmentation-based area measurements (LA area, RA area), 300 randomly sampled cases were manually segmented and used as the reference standard sonographer annotations and clinical LA area and RA area values are not available in MIMIC. For each parameter, (A) shows a scatter plot comparing three automated deep learning measurements (y-axis) with the reference LA area values (x-axis), (B) shows a scatter plot for RA area values. Detailed performance metrics are provided in **Supplemental Table 5**.

**Supplemental Methods.**

**YOLO model development for VTI peak detection.**

To localize the Doppler spectral envelopes on each VTI Doppler image, we trained a YOLOv11 object detector (Ultralytics). The YOLOv11-nano (yolo11n) was used for all reported models. All models were initialized from COCO-pretrained weights and fine-tuned on our Doppler dataset. Input images were resized to 640 pixels and models were trained for a maximum of 150 epochs with a batch size of 200, using early stopping with a patience of 10 epochs. The dataset was split into training, validation, and test splits by patient level. The checkpoint achieving the best validation performance was retained as the final model. Detection performance was evaluated on the held-out test split using IoU, mAP@50 and mAP@50-95.
